# Pharmacokinetics, bactericidal activity and toxicity of short oral regimens for rifampicin-resistant tuberculosis treatment

**DOI:** 10.64898/2026.03.24.26349145

**Authors:** Bern-Thomas Nyang’wa, Ilaria Motta, Ronelle Moodliar, Varvara Solodovnikova, Shakira Rajaram, Mohammed Rasool, Catherine Berry, David A.J. Moore, Geraint Davies, Frank Kloprogge

**Affiliations:** Médecins sans Frontieres, Public Health Department, Amsterdam, The Netherlands; University College London, MRC-CTU, London, United Kingdom; THINK (TB & HIV Investigative Network) Durban, South Africa; Republican Scientific and Practical Centre of Pulmonology and Tuberculosis, Minsk, Belarus; Clinical HIV Research Unit (CHRU), Wits Health Consortium (WHC), Department of Internal Medicine, School of Clinical medicine, Faculty of Health Sciences, University of Witwatersrand, Johannesburg, South Africa; Médecins sans Frontieres, Manson Unit, Public Health Department, London, United Kingdom; London School of Hygiene and Tropical Medicine, Clinical Research Department, London, United Kingdom; University of Liverpool, Department of Clinical Infection, Microbiology and Immunology, Liverpool, United Kingdom; Institute for Global Health, University College London, London, United Kingdom

**Author notes:** Joint senior authors (Geraint Davies, Frank Kloprogge). **Corresponding authors:** Bern-Thomas Nyang’wa; Frank Kloprogge.

## Abstract

WHO recommends bedaquiline-pretomanid-linezolid-(BPaL) and BPaL-moxifloxacin (BPaLM) for treatment of rifampicin-resistant tuberculosis, informed by the TB-PRACTECAL results. However, clinical explanatory data of these drugs’ exposure and *Mycobacterium tuberculosis* clearance rates and toxicity relationships remain understudied. We therefore investigated the relationship between the patients’ exposure to anti-TB drugs in TB-PRACTECAL trial investigational regimens and their treatment outcomes.

PRACTECAL-PKPD was a prospective pharmacokinetics and pharmacodynamics study nested in TB-PRACTECAL. Patients with rifampicin-resistant pulmonary tuberculosis were enrolled from Belarus and South Africa. The first objective was to develop drug exposure metrics for bedaquiline, pretomanid, linezolid, moxifloxacin and clofazimine. The efficacy objectives were to establish an exposure-response model for each drug and regimen to both bactericidal activity and long-term treatment outcomes. The safety objective was to investigate the exposure-toxicity relationship of each drug.

Antimicrobial exposure did not correlate with the speed of sputum bacterial clearance, however there was a 20% increased bacillary killing rate with BPaLM compared to the standard of care arm whilst BPaL and BPaL-clofazimine (BPaLC) displayed a 15% decreased bacillary killing rate compared to the standard of care arm. Linezolid plasma exposure was higher amongst patients with anaemia or neutropenia compared to those without. No other exposure-toxicity relationships were identified for all other drugs.

Absence of correlation between drug exposure and bacillary clearance suggest that the dosages used achieve saturation of bacillary killing, while remaining safe.

The study was registered in Clinical Trials.gov, TRN: NCT04081077.

**Funding:** Médecins Sans Frontières.

## Introduction

Each year around 500,000 patients fall ill with tuberculosis that is resistant to rifampicin with or without additional resistance to isoniazid^1^. Until 2022, when WHO provisionally recommended the use of Bedaquiline-Pretomanid-Linezolid-Moxifloxacin (BPaLM) for treatment of drug-resistant TB, treatment was long at 9-20 months with considerable toxicity and poor efficacy at around 60%^2^. Despite the Bedaquiline-Pretomanid-Linezolid (BPaL) backbone showing superior efficacy compared to local standard of care (SoC) regimens, at reduced treatment duration with less toxicity, *Mycobacterium tuberculosis* (*Mtb*) clearance from sputum and toxicity and their correlation with antimicrobial exposure remains understudied. A detailed understanding could facilitate further dose optimisation to maximise bactericidal activity whilst minimising toxicity.

Bedaquiline is a diarylquinoline antimycobacterial which inhibits the proton pump of mycobacterial ATP synthase ^3^. Pretomanid is a nitroimidazooxazine that inhibits mycolic acid biosynthesis, thus disrupting cell wall production in actively replicating Mtb ^4^; Linezolid is an oxazolidinone that inhibits ribosomal 8 protein synthesis by binding to the 23S RNA peptidyl transferase centre of the 50S subunit of the prokaryotic ribosome ^5^; Clofazimine is a lipophilic riminophenazine that acts through intracellular redox cycling, interfering with potassium uptake in membrane phospholipids and anti-inflammatory activity through inhibition of T-lymphocytes activation and proliferation^6^. Moxifloxacin is a fluoroquinolone that inhibits DNA gyrase that ultimately inhibits replication of M.tb DNA^7^.

Linezolid is part of the investigational treatment backbone in TB-PRACTECAL although there are concerns around safety of the administered dose. The most widely used dose of 600 mg is known to be associated with toxicity given the long duration of linezolid intake for treatment of *Mtb*. Dose reduction to 300 mg is therefore not uncommon and was used routinely after four months in TB-PRACTECAL ^8,9^. All other drugs were given at the labelled doses.

Pharmacokinetic–pharmacodynamic (PK–PD) indices, describing the association between antibiotic exposure and *Mtb* clearance have been derived from animal models or hollow-fiber systems for tuberculosis (HFS-TB), typically under monotherapy conditions ^10–13^. Subsequent probability of target attainment (PTA) simulations, that have informed dosing regimens, may consequently indicate suboptimal target attainment as the drug combination effect is being accounted for. However, assuming at least additive effects, exposures may still be sufficient when drugs are administered in combination to achieve complete eradication of *Mtb*.

Sputum *Mtb* load data from patients receiving combination treatment is available but is typically analysed as binary variable, i.e. conversion to negative *Mtb* sputum after two or three months of treatment ^14^. This approach omits data and therefore discards information. Characterising longitudinal *Mtb* sputum load data using statistical modelling techniques enables summarising how sputum clearance evolves over time and enables correlating longitudinal changes in sputum *Mtb* load with treatment efficacy ^15,16^.

We aimed to study the relationship between the patients’ exposure to anti-TB drugs in the TB-PRACTECAL trial investigational regimens and their treatment outcomes.

## Results

Between January 2017 and March 2021, 552 patients were enrolled in the main study, of whom 94, 458 and 514 patients contributed to the pharmacokinetic, mycobacteriology and adverse event analyses. The baseline age and BMI of all three populations were similar, however the proportion with HIV was higher at 42% for the PK population. 359 patients contributed two or more MGIT-TTP data samples during the first 12 weeks treatment as a measure of sputum bacillary load evolution over time, 274 (76%) had smear-positive tuberculosis, and 256 (71%) had tuberculosis cavities (Table 1).

**Table 1:** Baseline characteristics of the study population.

|  | Pharmacokinetics | Mycobacteriology | Toxicity | TB-PRACTECAL |
| --- | --- | --- | --- | --- |
| **Patients (n)** | 94 | 359 | 514 | 552 |
| **Age (years, median [IQR])** | 36 [31-43] | 35 [27-43] | 35 [27-43] | 35 [27-43] |
| **Female (n,%)** | 34 [36] | 147 [41] | 216 [42] | 223 [40] |
| **Weight (kg, [IQR])** | 56.8 [48.2-67.775] | 55.9 [48.5-64.05] | 56.5 [49.925-65] | 56.9 [50-65.625] |
| **BMI (kg/m<sup>2</sup>, [IQR])** | 19.75 [17.6-22.2] | 19.2 [17.45-22] | 19.8 [17.6-22.3] | 19.7 [17.7-22.3] |
| **HIV (n,[%])** | 39 [41] | 108 [30] | 146 [28] | 153 [28] |
| **Smear 1+ (n,[%])** | 9 [10] | 57 [16] | 69 [13] | 75 [14] |
| **Smear 2+ (n,[%])** | 12 [13] | 105 [29] | 110 [21] | 114 [21] |
| **Smear 3+ (n,[%])** | 29 [31] | 112 [31] | 110 [21] | 118 [21] |
| **Cavity (n,[%])** | 37 [40] | 256 [71] | 310 [60] | 326 [59] |

IQR: inter-quartile range, BMI: body mass index

Antimicrobial drug exposure was available for 57 patients contributing two or more MGIT-TTP data, while all 94 patients had both drug exposure and related adverse event data (Figure 1 & Table 1). Study profile per arm is in appendix figure S1.

**Figure 1:**
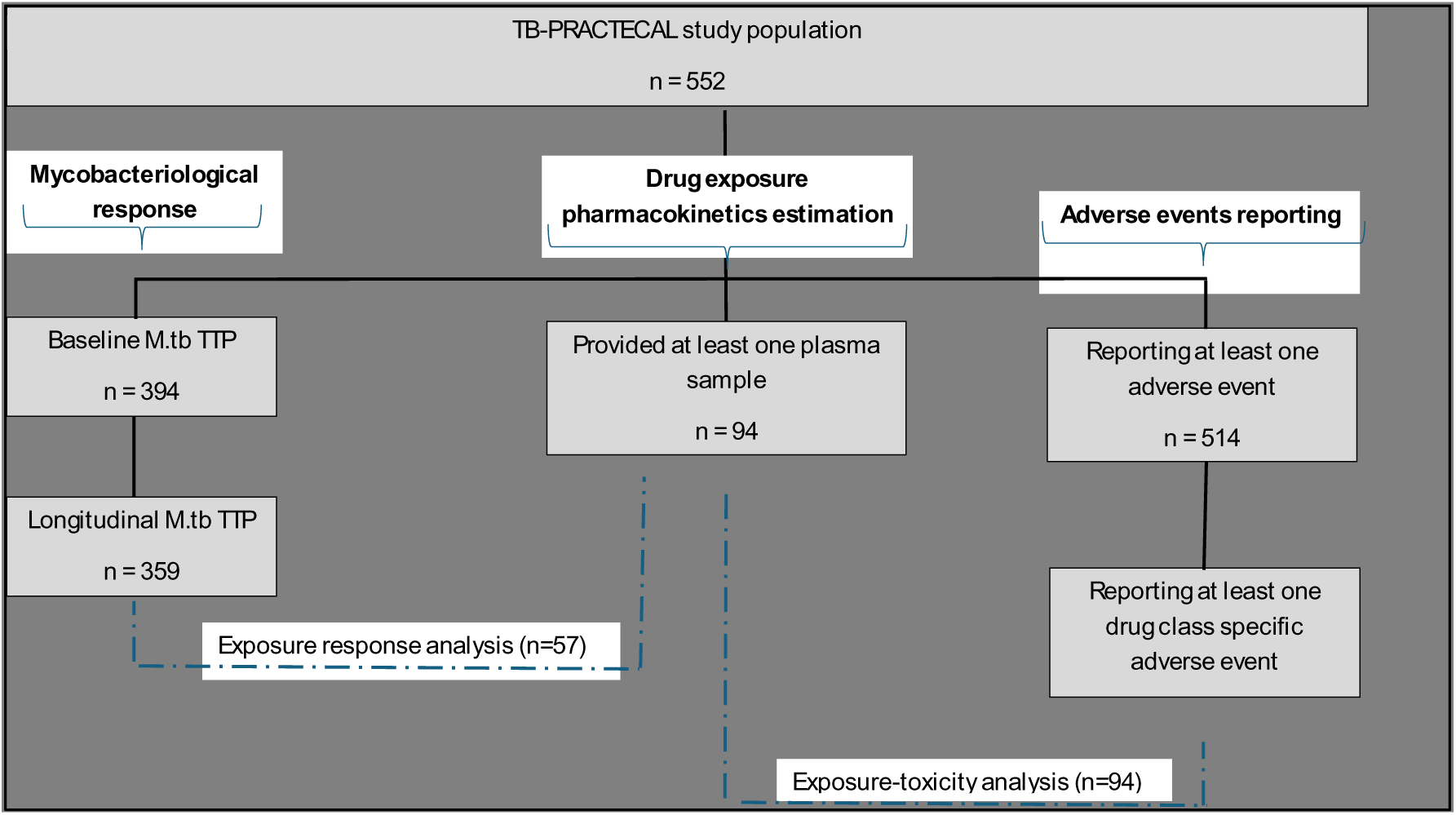
Study profile and analyses populations.

For 37 patients in the BPaLM arm, 298 (median [range], 9 [4-9] samples/patient) moxifloxacin plasma concentration samples were available. Absorption, distribution and elimination of moxifloxacin in plasma was described using a two-compartment distribution model, first-order absorption and elimination (Table S1 and Figure S2). Elimination clearance was at 14.2 l/h (%CV, 23.1) and corresponding median [IQR] steady state moxifloxacin AUC_0-24_ was 26.5 [20.0-40.0] mg*hr/l (Table 2). Across the BPaL, BPaLC and BPaLM arms, a total of 94 patients were enrolled that contributed 952 (median [range], 11 [3 - 12] samples/patient) bedaquiline plasma concentration samples. Absorption, distribution and elimination of bedaquiline in plasma was described using a three-compartment distribution model, transit absorption and elimination (Table S1 and Figure S3). Elimination clearance was at 1.93 l/h (%CV, 43.1) and corresponding median [range] bedaquiline AUC_0-48_ and AUC_0-72_ was 74.3 [24.6-140] and 104.5 [32.9-201] mg*hr/l at treatment completion. Exposure estimates for all study drugs are summarised in table 2. Population PK model estimates for pretomanid, linezolid and clofazimine which have been previously reported^17–19^, and for both moxifloxacin and bedaquiline are summarised in Table S1.

**Table 2:**
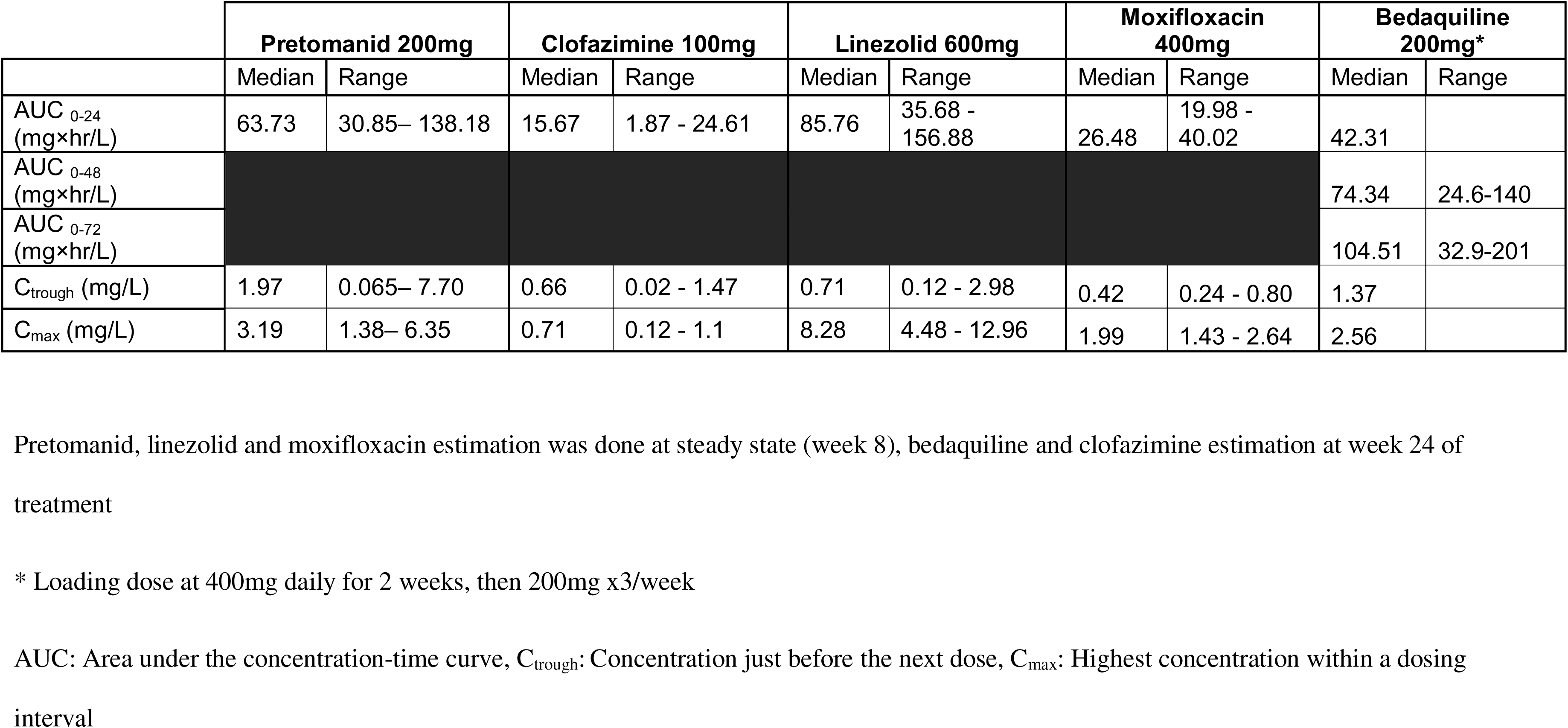
Individual drug exposure metrics.

Baseline sputum bacillary load expressed as the median MGIT time to positivity (MGIT TTP) in days was 14.4, 8.9, 6.2 and 5.1 days for patients with smear negative, 1+, 2+, and 3+ respectively. The bacillary clearance model VPCs demonstrated their predictive accuracy (Figure S4).

Sputum bacillary clearance was slower for patients with smear 2+ at baseline, at a MGIT TTP slope of 0.804 compared to smear 1+ or negative patients. Cavitary disease at baseline also resulted in reduced sputum bacillary clearance at a MGIT TTP slope of 0.728 compared to cavitary negative patients. Sputum bacillary clearance in the BPaLM arm was faster compared to the SoC arm whilst the sputum bacillary clearance in the BPaL/BPaLC arm was slower compared to the SoC arm. The MGIT-TTP slope for the BPaLM arm was 20% steeper than SoC arm whilst MGIT-TTP slope in the BPaL/BPaLC arm was 15% less steep than the SoC. (Figure 2 and Table S2).

**Figure 2:**
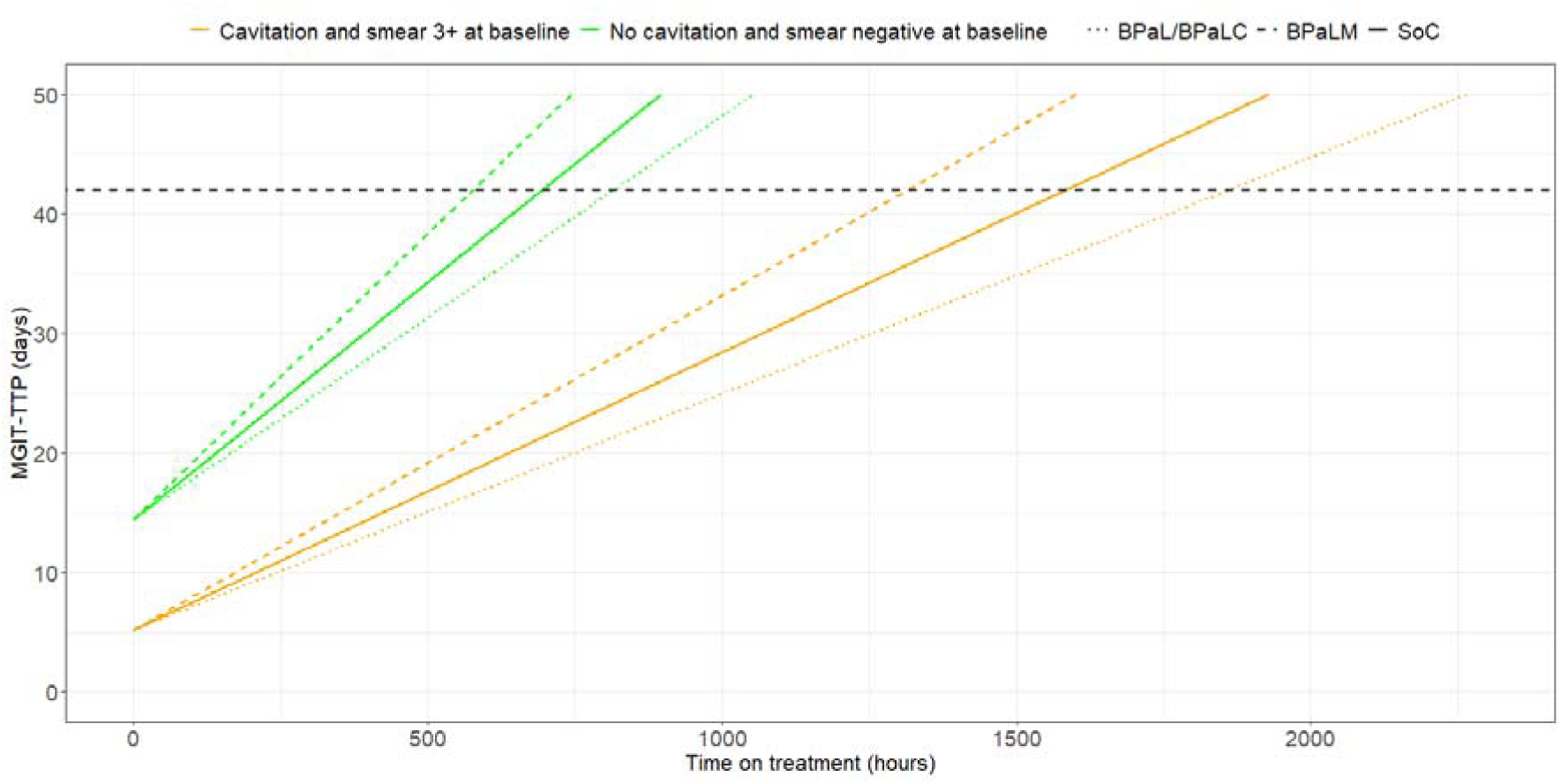
Effect of baseline covariates and treatment regimens on bacillary clearance. Using model-predicted MGIT-TTP vs time on treatment profiles for six representative patients. The participants with favourable baseline covariates (negative smear and absence of lung cavitation) are shown in green, whereas the participants with unfavourable baseline covariates (smear 3+ and presence of lung cavitation) are shown in orange. The solid, dotted, and dot-dashed lines represent SoC, BPaL/BPaLC, and BPaLM, respectively. The black dashed horizontal line represents a TTP of 42 days. MGIT-TTP, time-to-positivity. SoC: Standard of Care, BPaL/BPaLC: bedaquiline, pretomanid, linezolid plus/minus clofazimine, and BPaLM: bedaquiline, pretomanid, linezolid and moxifloxacin.

Neither antimicrobial bedaquiline, moxifloxacin or previously reported pretomanid, linezolid, clofazimine exposures correlated with random variability on sputum bacterial clearance between patients in EBE plots (figure 3) and when explored as a fixed effect covariate. And there were no differences in the antimicrobial exposure by the various long term trial outcomes (figure S5).

**Figure 3a:**
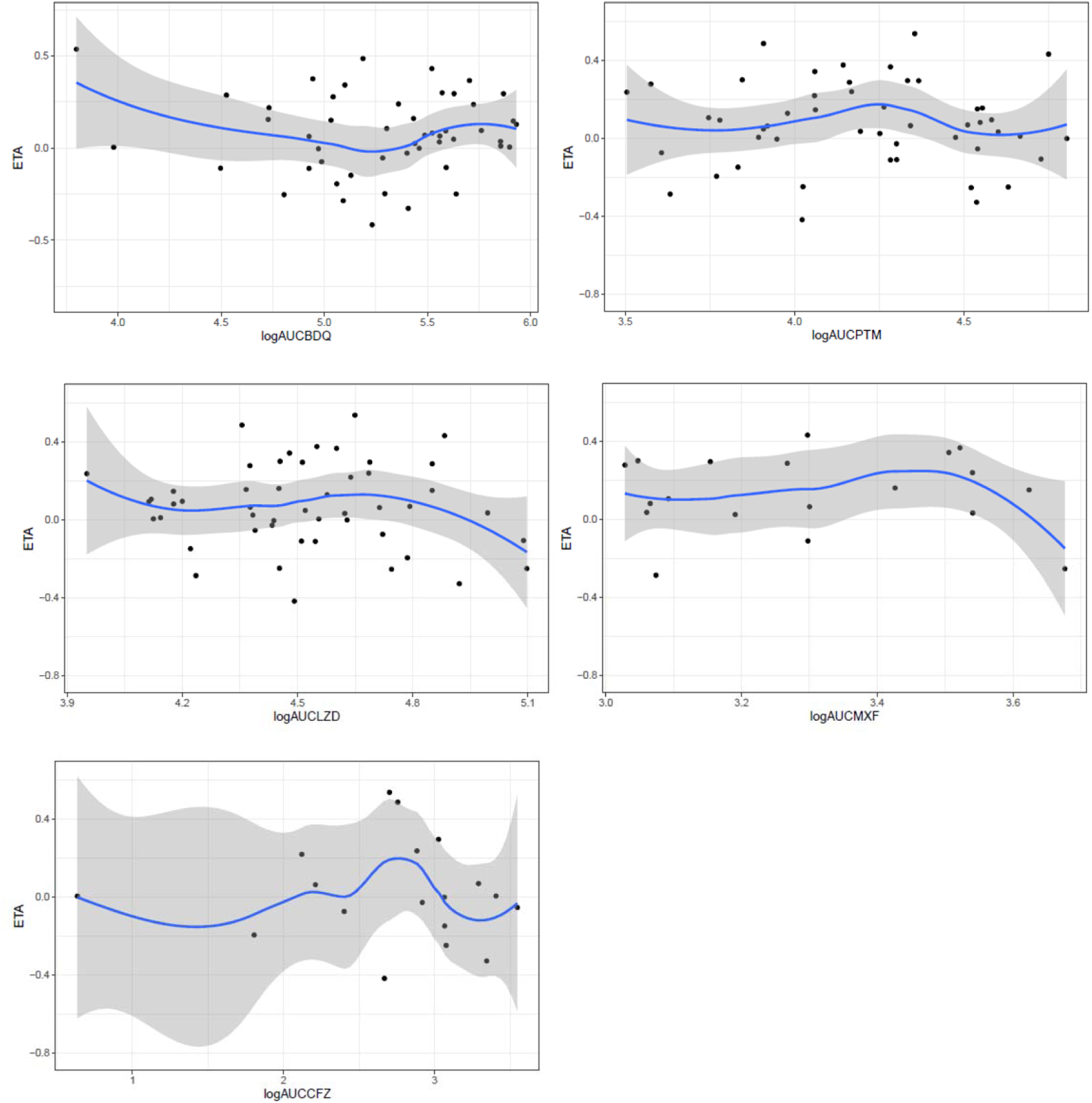
Individual drug exposure-response plots of random variability on TTP (ETA) slope vs individual model predicted log AUC_0-24_ for each of the study drugs.

**Figure 3b:**
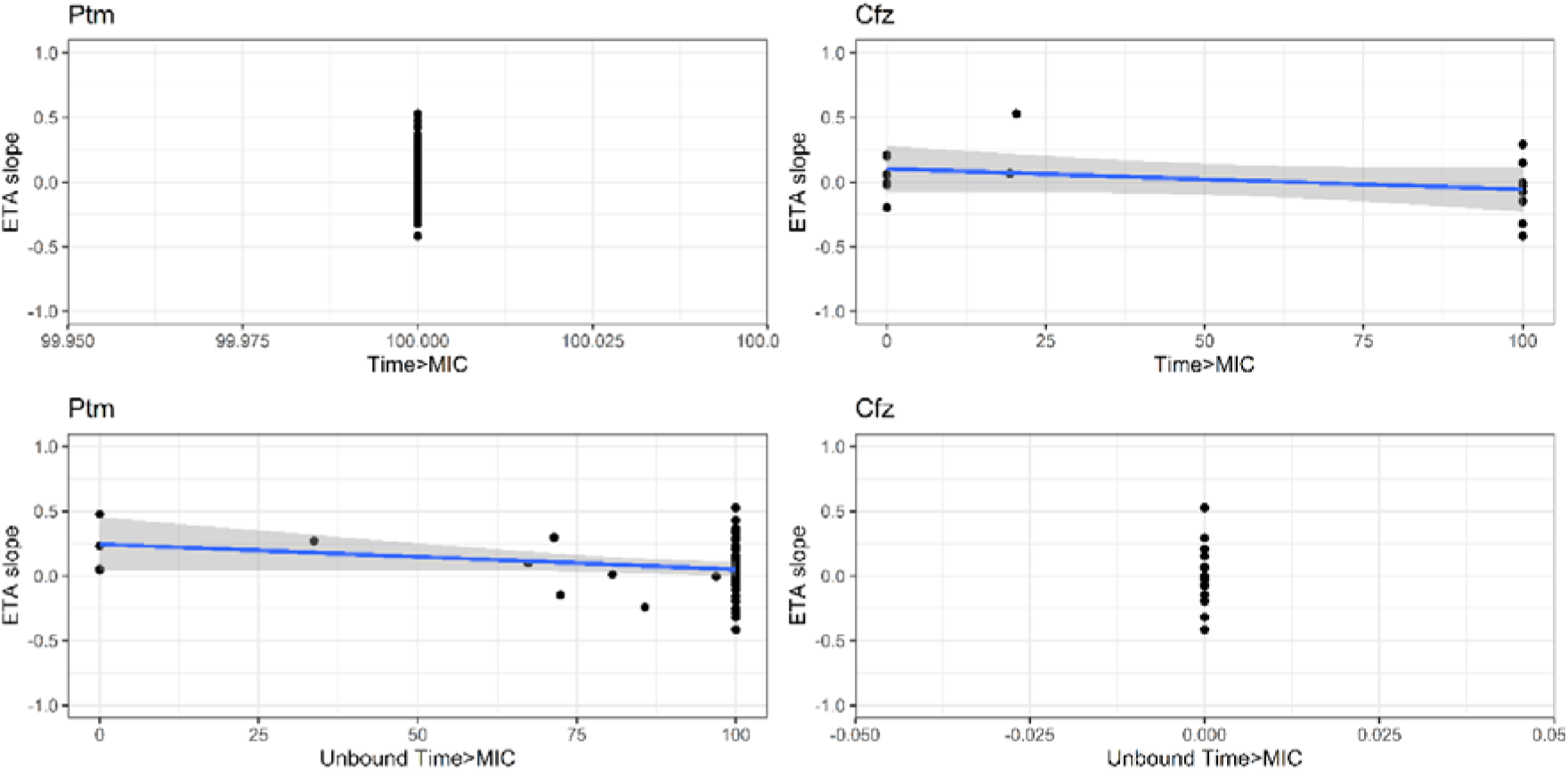
Individual drug exposure-response plots of random variability on TTP (ETA) slope vs individual model predicted T>MIC for pretomanid and clofazimine. The blue line with corresponding standard error represents the loess.

A total of 280 patients had anaemia, decreased creatinine clearance, elevated ALT, AST, bilirubin, neutropenia or peripheral neuropathy, ranging from grade 1-4, 89, 55, 65, and 71 patients in the SoC, BPaL, BPaLC and BPaLM arm (Figure S5). For those with antimicrobial plasma concentrations, 38 patients had AEs, ranging between grade 1 and grade 2. In one way ANOVA analyses, neither bedaquiline, pretomanid, moxifloxacin nor clofazimine AUC_0-24_ was correlated with liver dysfunction, i.e. ≥grade 1 elevated AST, ALT or bilirubin. Linezolid AUC_0-24_ was correlated with ≥grade 1 anaemia and neutropenia (Figure 4). Individual bedaquiline, pretomanid, linezolid moxifloxacin or clofazimine AUC_0-24_ did not correlate with grade for each of the investigated adverse events, with most of the adverse events being grade 1.

**Figure 4:**
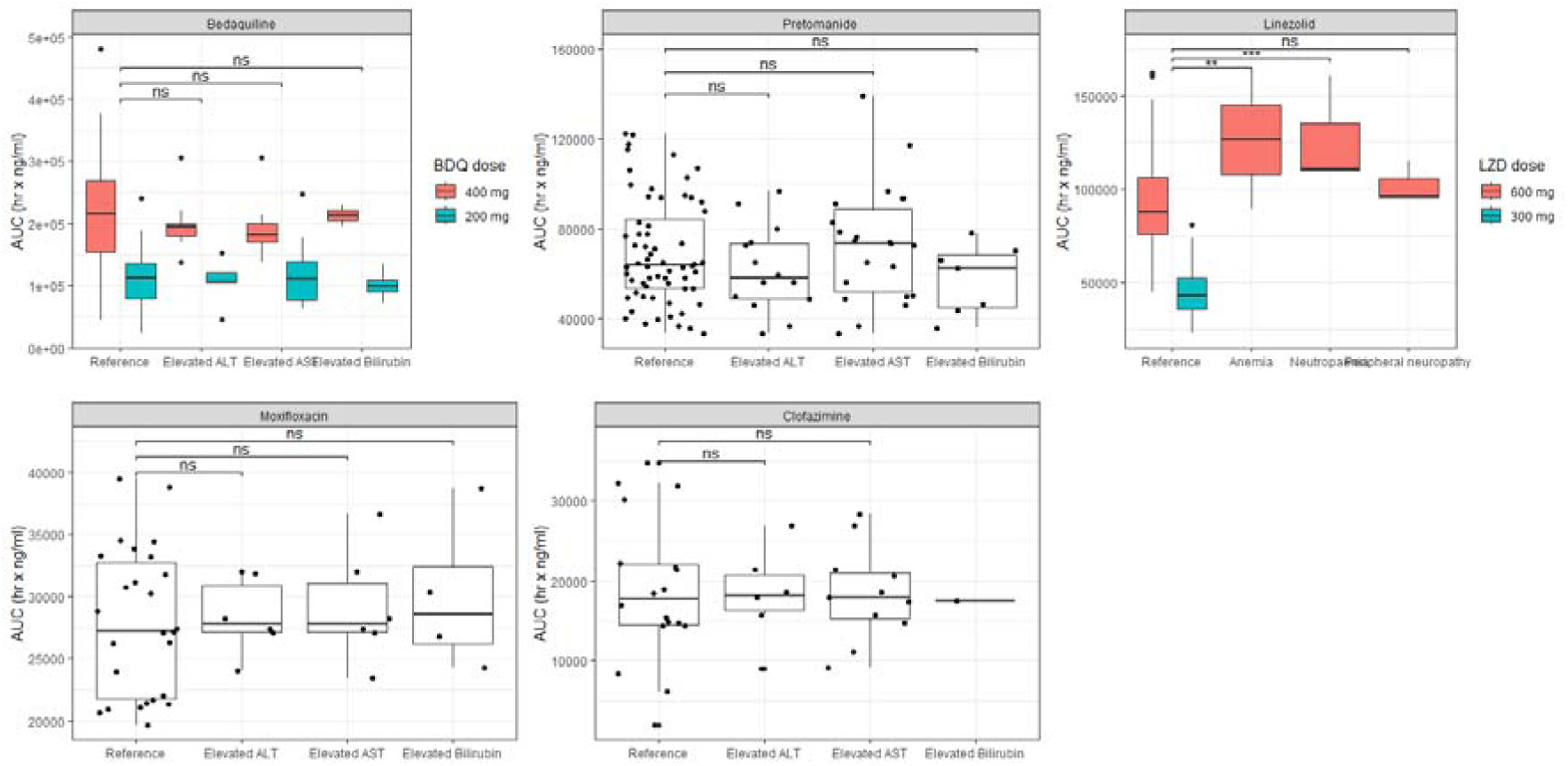
Box plots of AUC_0-24_ distribution of bedaquiline, pretomanid, linezolid, moxifloxacin and clofazimine for each related adverse event. One way ANOVA results: ns (no difference), ** p value of <0.01, *** p value of <0.001.

## Discussion

This study investigated plasma antibiotic exposure of participants in the TB-PRACTECAL trial, and its relationship with *Mtb* sputum clearance rates, efficacy and safety outcomes.

Moxifloxacin population pharmacokinetic properties aligned with published work, elimination clearance of 14.2 l/hr is within the range of previously reported estimates at 8-18 l/hr (*6, 20, 21*). Bedaquiline disposition kinetics, three-compartments, and elimination clearance at 1.93 l/hr, corresponded with previously reported population multi-compartmental pharmacokinetic models reporting elimination clearances at 1.50 – 4.54 l/hr^3,11,20–22^. Plasma exposures for bedaquiline, pretomanid, linezolid, moxifloxacin and clofazimine were within previously published efficacious ranges^17–19^.

Longitudinal data analyses of sputum *Mtb* load confirmed BPaLM’s superior capacity to clear *Mtb* from sputum compared to SoC, which was previously described by time to sputum conversion. The study also showed that BPaL and BPaLC were inferior in clearing *Mtb* from sputum compared to SoC, likely due to the contribution of the fluoroquinolone (97%), linezolid (87%) and bedaquiline (88%) in the SoC regimens. Amongst the investigated regimens in TB-PRACTECAL, BPaLM displayed highest bacillary clearance, and effectiveness on the primary endpoint of the trial while the inferior bacillary clearance of BPaL and BPaLC were also reflected in slightly higher rates of microbiological treatment failure and relapse ^9^. Although our findings show concordance between sputum bacillary clearance rates and treatment efficacy, this association may be context-specific and should not be interpreted as a universally predictive marker^23^.

Variability in rifampicin exposure after standard dosing has been well known to correlate with sputum *Mtb* clearance during first-line combination therapy of TB, highlighting that the pharmacodynamic effect is not saturated, supporting endeavours to increase the dose of rifampicin^15,24^. In this study, associations between *Mtb* sputum clearance and plasma antibiotic exposure were absent, which may argue that current doses achieve plasma levels sufficiently high enough to saturate *Mtb* clearance from sputum. However, the PK-PD dataset is small and may not have had sufficient power to detect differences among drugs with correlated exposures in the context of combination therapy with fixed dose levels. Furthermore, highly bactericidal drugs such as moxifloxacin may have similar effects to isoniazid, in masking the dose-response effect of accompanying drugs^25^.

A trend of higher linezolid AUC_0-24_ was observed in patients with anaemia, neurotoxicity and peripheral neuropathy compared to those patients without. However, the linezolid containing investigational arms in the TB-PRACTECAL study were generally well tolerated, with only grade 1 and 2 adverse events being observed amongst patients that also had linezolid concentrations quantified in plasma. This confirms previously reported associations between linezolid plasma exposure and toxicity, while it also underpins the success of a step down from daily linezolid at 600 mg to 300 mg after 16 weeks, to prevent toxicity^8^.

Though PRACTECAL-PKPD was not a dose-ranging trial, these data on early *Mtb* clearance from sputum and adverse events, taken in conjunction with the primary analysis, indicate that the studied doses of linezolid and moxifloxacin are effective and safe. This strengthens the WHO recommendation to include BPaLM as the preferred drug combination for treatment of pulmonary rifampicin-resistant tuberculosis in adolescents and adults.

## Methods

### Study design

PRACTECAL-PKPD was a prospective optimal design-based pharmacokinetics and pharmacodynamics study nested in TB-PRACTECAL - an open-label, randomised, controlled, multi-arm, phase IIb-III, multicentre, non-inferiority trial which investigated Bedaquiline-Pretomanid-Linezolid (BPaL), BPaL-Clofazimine (BPaLC), and BPaL-Moxifloxacin (BPaLM) compared to standard of care for patients with rifampicin-resistant pulmonary tuberculosis. Details of the protocols for both TB-PRACTECAL (<u>NCT02589782</u>), and PRACTECAL-PKPD (NCT04081077) have been previously published^26,27^.

Ethical approval was obtained from the London School of Hygiene & Tropical Medicine Research Ethics Committee and the Médecins Sans Frontières Ethics Review Board. Site specific approvals were obtained from the Belarus Republican Scientific and Practical Centre of Pulmonology and Tuberculosis ethics committee for the Minsk site, PharmaEthics for the Don Mckenzie and Dorris Goodwin hospitals sites and the University of Witwatersrand Human Research ethics committee for the Helen Joseph and King DiniZulu Hospitals sites. Written informed consent was obtained from all participants before recruitment into the TB-PRACTECAL trial and sub study-specific consent was obtained before any data or sample collection for the PRACTECAL-PKPD. All clinical activities were conducted in accordance with International Council for Harmonisation’s (ICH) Good Clinical Practice and laboratory investigations were conducted in accordance with Good Clinical and laboratory Practice

### Participants

Patients aged 15 years or older with rifampicin-resistant pulmonary tuberculosis enrolled in the investigational arms of TB-PRACTECAL at five hospitals and community centres in Belarus, and South Africa were offered enrolment into the PKPD study.

Full inclusion and exclusion criteria have been reported previously ^26,27^. Of note was the inclusion of patients with HIV irrespective of CD4 count and exclusion of those who were pregnant, with alanine aminotransferase (ALT) concentration or an aspartate aminotransferase (AST) concentration higher than three times the upper limit of the normal range, any baseline laboratory value consistent with Grade 4 toxicity or a Fridericia-corrected QT (QTcF) interval longer than 450 ms.

### Procedures

BPaLM, BPaLC and BPaL were taken orally for 24 weeks. Bedaquiline was administered at 400 mg daily for two weeks followed by 200 mg three times weekly for the remaining 22 weeks, pretomanid 200 mg daily for 24 weeks, and linezolid 600 mg daily for the first 16 weeks followed by 300 mg for the remaining 8 weeks. BPaLM additionally contained moxifloxacin 400 mg daily for 24 weeks and BPaLC, clofazimine 100 mg daily (or 50 mg for patients weighing <33kg) for 24 weeks. Comparator treatment comprised locally accepted standard-of-care (SoC), that was updated in line with international recommendations over the course of the study. Details of medication administration have been published previously ^27,28^.

For the pharmacokinetic analyses, plasma samples were collected at zero, two and 23 hours post dose during the baseline visit, at zero, 6.5 and 23 hours post dose during the week 8 visit, followed by trough samples at the week 12, 16, 20, and 24 visits and additional samples at week 32 and week 72. Drugs were quantified using high-performance liquid chromatography-tandem mass spectrometry^26^.

Participants submitted spot and early morning sputum samples at day 0, day 7 and weeks 4, 8, 12, 16, 20 and 24 during treatment for microbiological analyses. Time to positivity in primary culture and minimum inhibitory concentrations were determined in the BD BACTEC™ Mycobacterial Growth Indicator Tube system (MGIT™) (Becton & Dickinson (BD), details have been previously published ^29^.

Safety monitoring was conducted at least every 4 weeks and included blood chemistry analysis and physical examinations ^27^.

### Outcomes

The first objective was to develop drug exposure metrics for bedaquiline, pretomanid, linezolid, moxifloxacin and clofazimine. The key model derived outcomes were the area under the drug concentration curve (AUC), the lowest drug concentration just before the next dose (Ctrough), the highest drug concentration between two doses (Cmax). Together with cumulative percentage of the dosing interval that the drug concentration exceeds the MIC (T>MIC) drug exposure matrices were evaluated, adjusted for MIC or not, in the context of bacillary clearance.

The efficacy (second and third) objectives were to establish an exposure-response model for each drug and regimen to both early bactericidal activity and long-term treatment outcomes. The main endpoint was the change over time to the duration it took for a culture to have growth detected in MGIT, i.e. time to positivity (TTP).

The safety (fourth) objective was to investigate the exposure-toxicity relationship of each drug. The outcome measures for this analysis were adverse events, including elevated ALT, AST or bilirubin levels, decreased creatinine clearance, anaemia, neutropenia and peripheral neuropathy.

### Modelling Software and Methods

Analyses including nonlinear mixed-effect modelling were performed in R v4.1.2 using nlmixr2 for the population PK and pharmacodynamic model development.

Bedaquiline, pretomanid, linezolid and clofazimine AUC_0-24_ have been previously published ^17–19^, whilst moxifloxacin and bedaquiline AUC_0-24_ were derived from a moxifloxacin and bedaquiline population pharmacokinetic models described here. Moxifloxacin and bedaquiline absorption, distribution, metabolism and excretion were characterised using a population pharmacokinetic model. Due to a sparse pharmacokinetic sampling design, fixed-effects for absorption rate constant (ka), inter-compartmental clearance (q), and peripheral volume of distribution (Vp) for moxifloxacin were fixed to literature values whilst for bedaquiline only ka was fixed to literature values^12,20^. Fat Free Mass (FFM) and Body Mass Index (BMI) were embedded in the model a priori based on previous literature reports^20,30^. Antimicrobial exposure indices, i.e. Area Under the concentration-time Curve (AUC_0-24_)/MIC and T>MIC, were investigated as explanatory variables for variability on MGIT TTP slope over time.

MGIT TTP-TIME data from the first twelve weeks of treatment in patients contributing at least two or more MGIT TTP samples was retained for data analysis as later positive samples represented relapses. A linear, exponential and power equation with random effects representing between-patient and residual variability, were fitted to the data and compared for superiority using the Bayesian Information Criterion. MGIT-TTP values above limit of quantification were handled with the censoring method in nlmixr2, based on the M3 methods in NONMEM. The upper limit of quantification was set to 42 days in line with the threshold for a MGIT-TTP negative sample. Clinically relevant explanatory variables were tested as proportional covariates on baseline MGIT TTP and MGIT TTP slope over time (p < 0.01 for 1 dF); these included baseline cavitation, smear status, HIV status, age, and study arm and these were candidate explanatory variables for variability in baseline MGIT TTP and slope over time.

The relationship between elevated AST, ALT, and bilirubin, decreased creatinine clearance, anaemia, neutropenia, and peripheral neuropathy with AUC_0-24_ was analysed using one-way ANOVA. Severity grading 1, 2, 3, or 4, was based on the MSF severity grading scale, which is based on the standardised and commonly used toxicity table for infectious diseases, the Division of Microbiology and Infectious Diseases (DMID) grading system, complemented with a selection of terms from the NCI’s Common Terminology Criteria for Adverse Events (CTCAE) scale.

## Data Availability

Deidentified pharmacokinetic data will be available to researchers upon a written request to the Medical Director, Medecins sans Frontieres, Operational Centre Amsterdam, the Netherlands. Clinical and mycobacteriology data is available from TB-PACTS study ID 1035 at: https://c-path.org/tools-platforms/tb-pacts and World Health Organization Tuberculosis treatment individual patient data platform, University College London. Available at: https://www.ucl.ac.uk/global-health/research/z-research/tb-ipd-platform.

## Data sharing

Deidentified pharmacokinetic data will be available to researchers upon a written request to the Medical Director, Médecins sans Frontières, Operational Centre Amsterdam, the Netherlands. Clinical and mycobacteriology data is available from TB-PACTS study ID 1035 at: https://c-path.org/tools-platforms/tb-pacts and World Health Organization Tuberculosis treatment individual patient data platform, University College London. Available at: https://www.ucl.ac.uk/global-health/research/z-research/tb-ipd-platform.

## Contributions

Conception and design: B-TN, FK, GD and DAM. Data acquisition: IM, RM, VS, SR. PK modelling: BT-N and FK. First draft of manuscript: BT-N and FK. Revising and approval of manuscript: all

## Acknowledgments

FK is recipient of a Sir Henry Dale Fellowship jointly funded by the Wellcome Trust and the Royal Society (Grant Number 220587/Z/20/Z).

