## Supplement for "Pharmacokinetics, bactericidal activity and toxicity of short oral regimens for rifampicin-resistant tuberculosis treatment"

[#] Joint senior authors

### Corresponding authors

### Supplementary Appendix

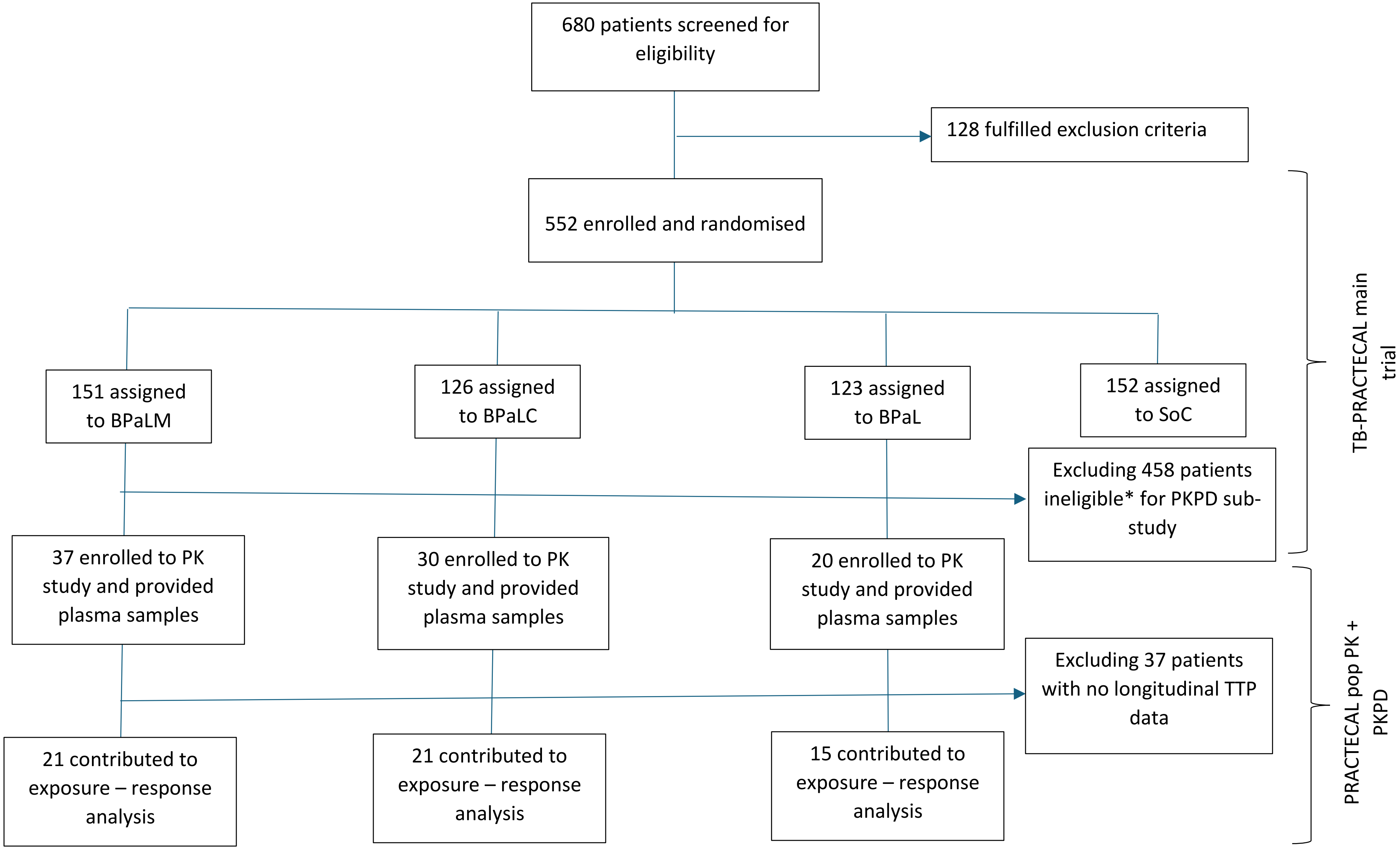


*ineligible = in SoC, at site not participating in sub-study, recruited into main trial before sub-study started

**Figure S1**: Study profile

**Pharmacokinetics**

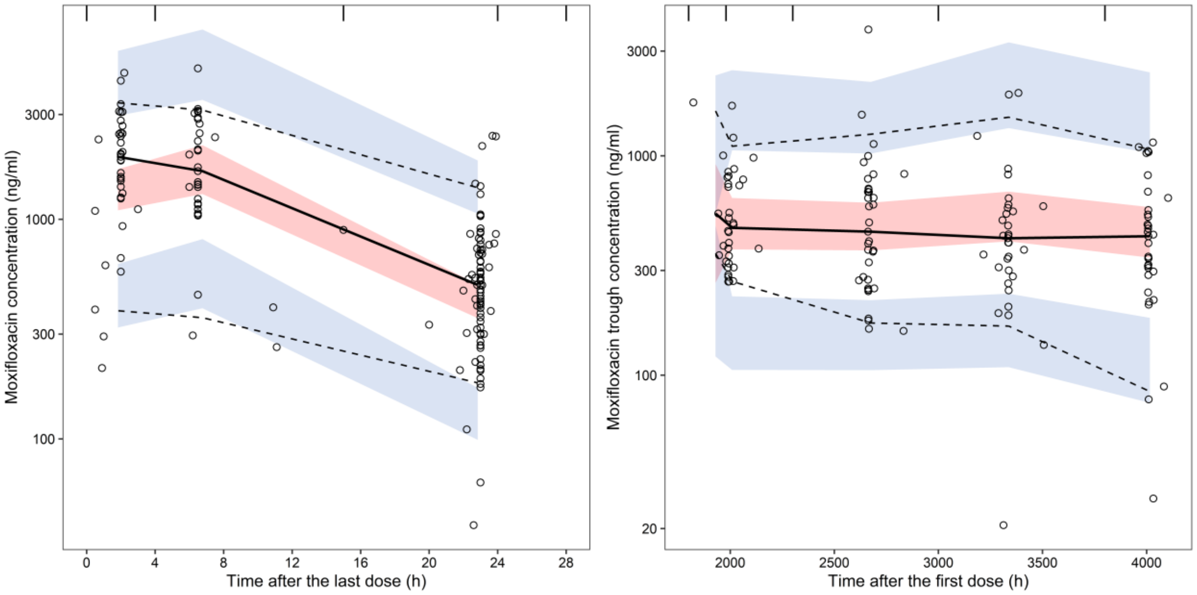


**Figure S2**: Visual predictive check of moxifloxacin plasma concentration vs time data. All data from the baseline and month two visit in the left panel as time after last dose, and through level data from the visits beyond month two in the right panel as time after the first dose. Dots represent observations, solid and dashed lines the median and 2.5-97.5 percentiles of the observe data with corresponding 90% Confidence Intervals overlaid ($n_{simulations}$ = 2,000).

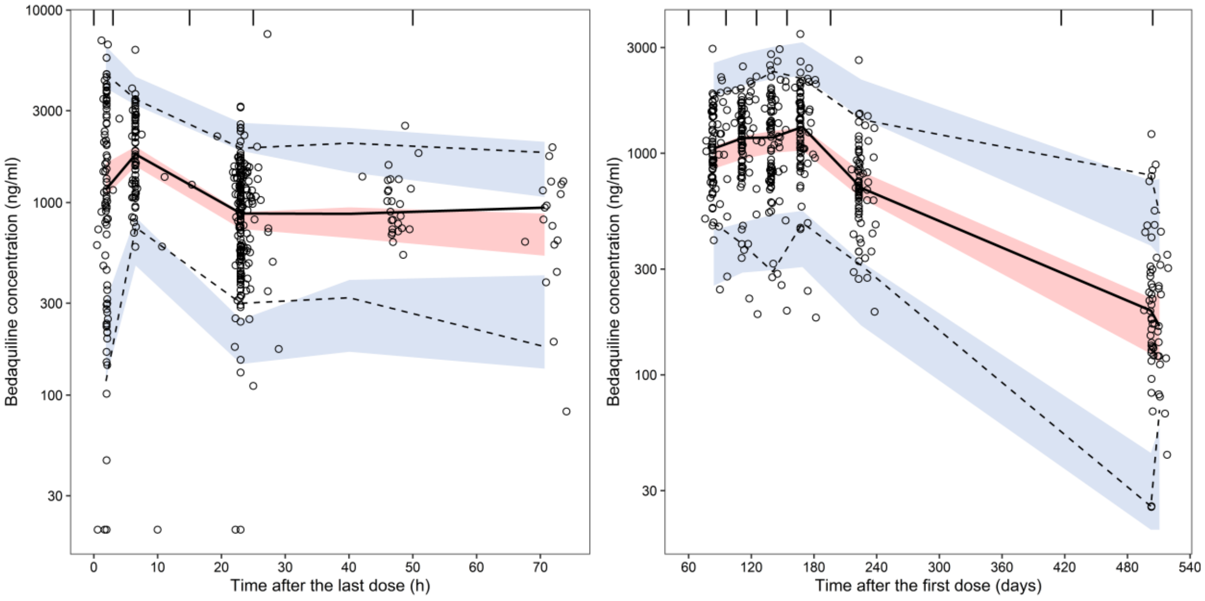


**Figure S3:** Visual predictive check of bedaquiline plasma concentration vs time data. All data from the baseline and month two visit in the left panel as time after last dose, and through level data from the visits beyond month two in the right panel as time after the first dose. Dots represent observations, solid and dashed lines the median and 2.5-97.5 percentiles of the observe data with corresponding 90% Confidence Intervals overlaid ($n_{simulations}$ = 2,000).

**Table S1:** Population pharmacokinetic model estimates for bedaquiline, pretomanid, linezolid, clofazimine and moxifloxacin of participants in the PRACTECAL-PKPD study.

| | Bedaquiline | | | | pretomanid | | | | linezolid | | | | clofazimine | | | | moxifloxacin | | | |
|---|---|---|---|---|---|---|---|---|---|---|---|---|---|---|---|---|---|---|---|---|
| **Parameter** | Mean estimate (95%CI) | %RSE | BSV (CV%) | Shrink % | Mean estimate (95%CI) | %RSE | BSV (CV%) | Shrink % | Mean estimate (95%CI) | %RSE | BSV (CV%) | Shrink % | Mean estimate (95%CI) | %RSE | BSV (CV%) | Shrink % | Mean estimate (95%CI) | %RSE | BSV (CV%) | Shrink % |
| **cl (l/h)** | 1.93 (1.58, 2.36) | 15.6 | 43.1 | 8.06%< | 3.1 (2.87, 3.33) | 3.36 | 32.9 | 9.65%< | 6.66 (6.04, 7.34) | 2.62 | 28.2 | 11.4%< | 6.84 (5.73, 8.15) | 4.68 | 77.3 | 8.78%< | 14.2 (12.2, 16.4) | 2.84 | 23.1 | 18.8%< |
| **vc (l)** | 103 (73.6, 143) | 3.65 | 132 | 10.3%< | 102 (81.4, 127) | 2.45 | 33.6 | 36.7%> | 58.8 (51.7, 66.9) | 1.61 | 5.63 | 94.6%> | 1.75e+03 (1.44e+03, 2.12e+03) | 1.32 | 173 | 2.39%< | 167 (133, 211) | 2.29 | 11.1 | 27.3%= |
| **vp (l)** | 6.51e+03 (5.45e+03, 7.77e+03) | 1.03 | 37.1 | 16.7%< | NA | NA | NA | NA | NA | NA | NA | NA | 9.15e+03 (7.79e+03, 1.07e+04) | 0.895 | 44.6 | 26.1%= | 87.4 | FIXED | | |
| **q (l/h)** | 7.49 (5.58, 10.1) | 7.45 | 66.1 | 15.7%< | NA | NA | NA | NA | NA | NA | NA | NA | 41.7 (34.6, 50.4) | 2.58 | 82 | 40.9%> | 2.3 | FIXED | | |
| **vp2 (l)** | 98.9 (49.3, 199) | 7.74 | | | NA | NA | NA | NA | NA | NA | NA | NA | NA | NA | NA | NA | NA | NA | NA | NA |
| **q2 (l/h)** | 7.02 (4.26, 11.6) | 13.1 | | | NA | NA | NA | NA | NA | NA | NA | NA | NA | NA | NA | NA | NA | NA | NA | NA |
| **prop.err** | 0.321 | | | | 0.322 | | | | NA | NA | NA | NA | 0.198 | | | | NA | NA | NA | NA |
| **add.err** | 59.5 | | | | 368 | | | | NA | NA | NA | NA | 16.4 | | | | NA | NA | NA | NA |
| **logn.err** | NA | NA | NA | NA | NA | NA | NA | NA | 0.888 | | | | NA | NA | NA | NA | 0.645 | | | |
| **ka (h-1)** | NA | NA | NA | NA | 0.316 (0.203, 0.492) | 19.6 | | | 1.23 | FIXED | | | 0.67 | FIXED | | | 0.569 | FIXED | | |
| **tlag (h)** | NA | NA | NA | NA | NA | NA | NA | NA | NA | NA | NA | NA | 0.62 | FIXED | | | NA | NA | NA | NA |
| **MAT** | 0.66 | FIXED | NA | NA | NA | NA | NA | NA | NA | NA | NA | NA | NA | NA | NA | NA | NA | NA | NA | NA |
| **FR** | 0.47 | FIXED | NA | NA | NA | NA | NA | NA | NA | NA | NA | NA | NA | NA | NA | NA | NA | NA | NA | NA |

BSV: between patient variability, Shrink: eta-shrinkage, RSE: relative standard error, prop.err: proportional error, add.err: additive error, logn.sd additive residual variability on both-sided log-transformed data. MAT refers to fraction of time for both delay and 90% complete absorption. Allometric body size scaling was applied to clearance(cl) and volume (v) parameters based on Fat Free Mass with the power fixed to 0.75 and 1, respectively.

**Efficacy outcomes**

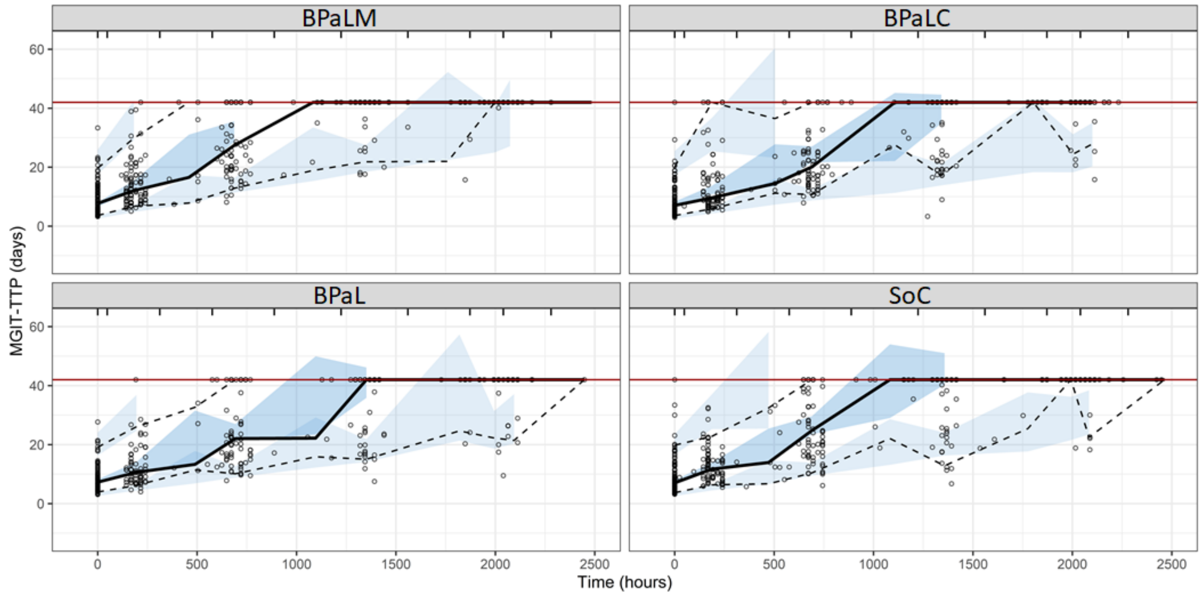


**Figure S4:** Visual predictive check of longitudinal MGIT-TTP in days vs time on treatment in hours, stratified by study arm. Dots represent observations, solid and dashed lines the median and 2.5-97.5 percentiles of the observe data with corresponding 90% Confidence Intervals overlaid ($n_{simulations}$ = 500). The horizontal red line represents the upper limit of quantification, i.e. 42 days).

**Table S2:** Population pharmacodynamic parameters for the MGIT-TTP model.

| | **Mean estimate(95%CI)** | **%RSE** | **BSV(CV%)** | **Shrink(SD)%** |
|---|---|---|---|---|
| **a** | 0.0397 (0.0341, 0.0463) | 2.43 | 30.7 | 24.4%= |
| **b** | 14.4 (13.2, 15.6) | 1.54 | 19.3 | 33.7%> |
| **ruv** | 0.393 | | | |
| **beta_b_Smear1** | 0.619 (0.552,0.693) | 12.1 | | |
| **beta_b_Smear2** | 0.431 (0.389,0.476) | 6.11 | | |
| **beta_b_Smear3** | 0.356 (0.319,0.398) | 5.47 | | |
| **beta_a_CAV** | 0.728 (0.642,0.825) | 20.1 | | |
| **beta_a_BPaLCMerged** | 0.852 (0.734,0.988) | 47.1 | | |
| **beta_a_BPaLM** | 1.204 (1.03,1.408) | 42.8 | | |
| **beta_a_Smear23** | 0.804 (0.712,0.908) | 28.5 | | |

BSV: between patient variability, Shrink: eta-shrinkage, RSE: relative standard error. ruv: residual unexplained variability

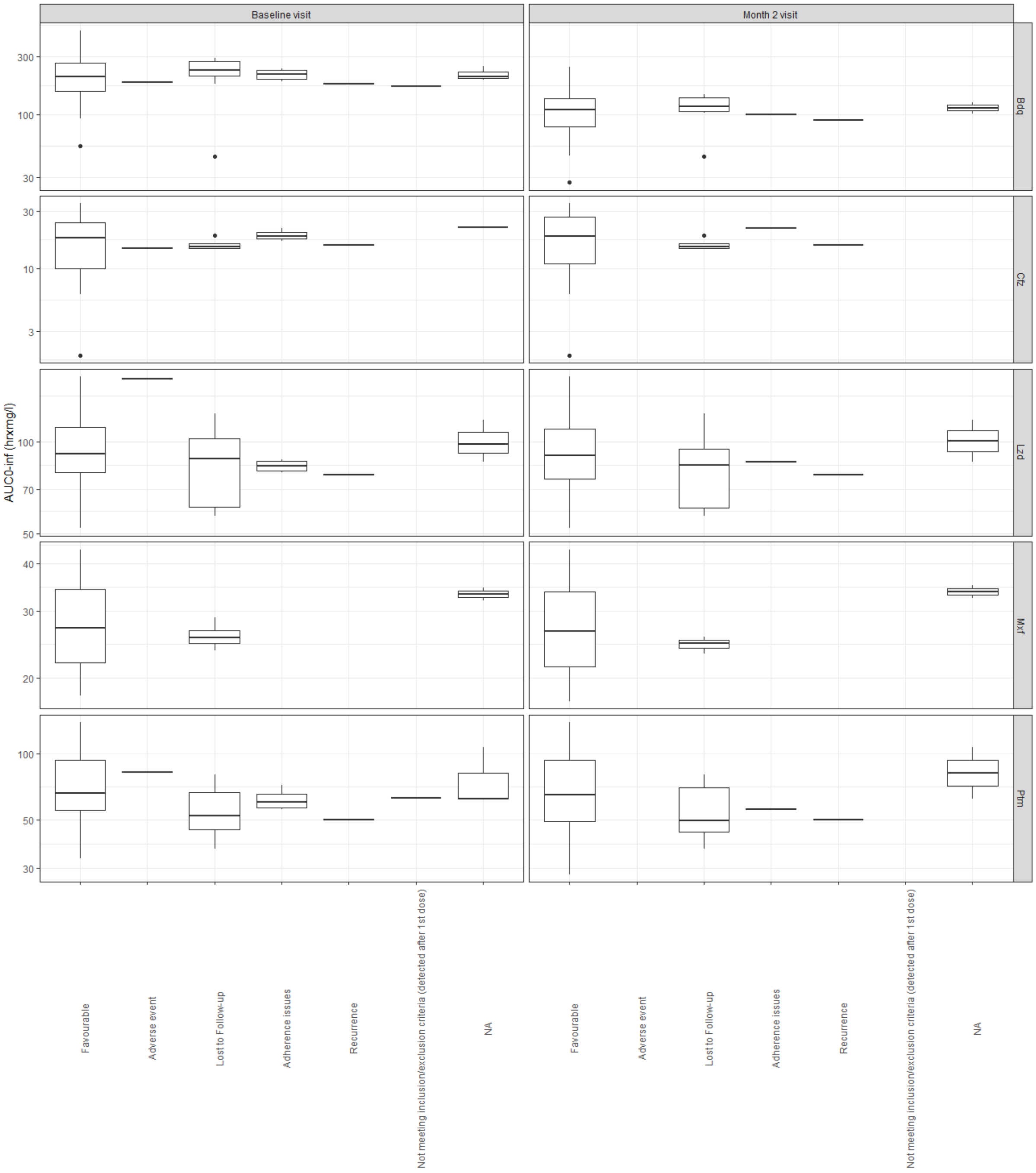


**Figure S5:** box plots of antimicrobial exposure by the various unsuccessful outcome categories.

### Safety outcomes

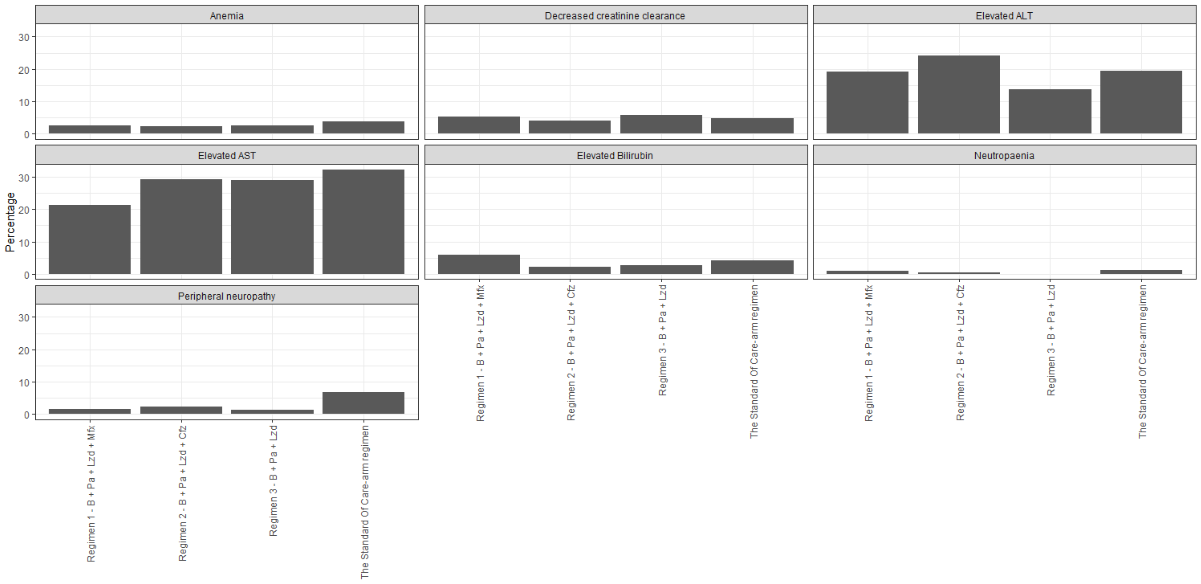


**Figure S6:** Visualisation of the occurrence of related adverse events per study arm.
